# The Causal Effects of Health Conditions and Risk Factors on Social and Socioeconomic Outcomes: Mendelian Randomization in UK Biobank

**DOI:** 10.1101/19008250

**Authors:** Sean Harrison, Alisha R Davies, Matt Dickson, Jessica Tyrrell, Michael J Green, Srinivasa Vittal Katikireddi, Desmond Campbell, Marcus Munafò, Padraig Dixon, Hayley E Jones, Frances Rice, Neil M Davies, Laura D Howe

## Abstract

**Objectives:** To estimate the causal effect of health conditions and risk factors on social and socioeconomic outcomes in UK Biobank. Evidence on socioeconomic impacts is important to understand because it can help governments, policy-makers and decision-makers allocate resources efficiently and effectively.

**Design:** We used Mendelian randomization to estimate the causal effects of eight health conditions (asthma, breast cancer, coronary heart disease, depression, eczema, migraine, osteoarthritis, type 2 diabetes) and five health risk factors (alcohol intake, body mass index [BMI], cholesterol, systolic blood pressure, smoking) on 19 social and socioeconomic outcomes.

**Setting:** UK Biobank.

**Participants:** 337,009 men and women of white British ancestry, aged between 39 and 72 years.

**Main outcome measures:** Annual household income, employment, deprivation (measured by the Townsend deprivation index [TDI]), degree level education, happiness, loneliness, and 13 other social and socioeconomic outcomes.

**Results:** Results suggested that BMI, smoking and alcohol intake affect many socioeconomic outcomes. For example, smoking was estimated to reduce household income (mean difference = −£24,394, 95% confidence interval (CI): −£33,403 to −£15,384), the chance of owning accommodation (absolute percentage change [APC] = −21.5%, 95% CI: −29.3% to −13.6%), being satisfied with health (APC = −32.4%, 95% CI: −48.9% to −15.8%), and of obtaining a university degree (APC = −73.8%, 95% CI: −90.7% to −56.9%), while also increasing deprivation (mean difference in TDI = 1.89, 95% CI: 1.13 to 2.64, approximately 236% of a decile of TDI). There was evidence that asthma increased deprivation and decreased both household income and the chance of obtaining a university degree, and migraine reduced the chance of having a weekly leisure or social activity, especially in men. For other associations, estimates were null.

**Conclusions:** Higher BMI, alcohol intake and smoking were all estimated to adversely affect multiple social and socioeconomic outcomes. Effects were not detected between health conditions and socioeconomic outcomes using Mendelian randomization, with the exceptions of depression, asthma and migraines. This may reflect true null associations, selection bias given the relative health and age of participants in UK Biobank, and/or lack of power to detect effects.

**What is known?:**

- Studies have shown associations between poor health and adverse social (e.g. wellbeing, social contact) and socioeconomic (e.g. educational attainment, income, employment) outcomes, but there is also strong evidence that social and socioeconomic factors influence health.
- These bidirectional relationships make it difficult to establish whether health conditions and health risk factors have causal effects on social and socioeconomic outcomes.
- Mendelian randomization is a technique that uses genetic variants robustly related to an exposure of interest (here, health conditions and risk factors for poor health) as a proxy for the exposure.
- Since genetic variants are randomly allocated at conception, they tend to be unrelated to the factors that typically confound observational studies, and are less likely to suffer from reverse causality, making causal inference from Mendelian randomization analyses more plausible.

**What this study adds:**

- This study suggests causal effects of higher BMI, smoking and alcohol use on a range of social and socioeconomic outcomes, implying that population-level improvements in these risk factors may, in addition to the well-known health benefits, have social and socioeconomic benefits for individuals and society.
- There was evidence that asthma increased deprivation, decreased household income and the chance of having a university degree, migraine reduced the chance of having a weekly leisure or social activity, especially in men, and depression increased loneliness and decreased happiness.
- There was little evidence for causal effects of cholesterol, systolic blood pressure or breast cancer on social and socioeconomic outcomes.

## 1. Background

Poor health has the potential to affect an individual’s ability to engage with society (1–4). For example, illnesses or adverse health behaviours could influence the ability to attend and concentrate at school or work and hence affect educational attainment, employment, and income. Illness and health behaviours may also affect an individual’s ability to maintain wellbeing and an active social life. From an individual perspective, maintaining good health can therefore have considerable social and socioeconomic benefits (5). Similarly from a population perspective, improving population health could lead to a happier and more productive population (6).

Understanding the causal impacts of health on social and socioeconomic outcomes can help demonstrate the potential broader benefits of investing in effective health policy, thereby strengthening the case for cross-governmental action to improve health and its wider determinants at the population-level (7). Furthermore, patients require accurate information about how their lives might be affected by their health, for example on returning to work after cancer (8). However, studying the social and socioeconomic consequences of ill health (‘social drift’) is challenging because of social causation, i.e. the strong role of social and socioeconomic circumstances in disease causation. Social causation means that associations between health and social and socioeconomic outcomes are likely to be severely biased by confounding and reverse causality. Methodological approaches strengthening causal inference in this field are therefore essential.

Mendelian randomization is a technique that uses genetic variants robustly related to an exposure of interest (here, health conditions and risk factors for poor health) as proxies for the exposure (instrumental variables) (9,10). Since genetic variants are randomly allocated at conception, results from Mendelian randomization studies are much less likely to suffer from confounding and reverse causality than traditional observational studies (11). In this paper, we apply Mendelian randomization within a large study of UK individuals aged between 39 and 72 years to estimate the causal effects of health conditions and risk factors with the greatest burden on UK adults on a range of social (e.g. social contact, wellbeing, and cohabitation status) and socioeconomic (e.g. education, employment, income) outcomes.

## 2. Methods

### Population

UK Biobank is a population-based health research resource consisting of approximately 500,000 people, who were recruited between the years 2006 and 2010 from 22 centres across the UK (12). Participants provided medical history and socioeconomic information via questionnaires, interviews and anthropometric measures at recruitment. Medical data from hospital episode statistics (HES) and the cancer registry have been linked to participants. The study design, participants and quality control methods have been described in detail previously (13–15). UK Biobank received ethics approval from the Research Ethics Committee (REC reference for UK Biobank is 11/NW/0382).

We restricted analyses to unrelated individuals of white British ancestry. Full details of inclusion criteria and genotyping are in **Supplementary Information 1**. After exclusions, 337,009 participants remained in the dataset.

### Measures of Health Conditions and Risk Factors (Exposures)

We used the Global Burden of Disease Study 2010 (GBD) (16) to identify health conditions and risk factors that contributed 100 or more disability-adjusted life years lost per 100,000 adults in the UK. From this list, we restricted our analysis to health conditions and risk factors with known genetic determinants and a prevalence of ≥2% among UK Biobank participants. This resulted in the inclusion of eight health conditions: asthma, breast cancer, coronary heart disease, depression, eczema, migraine, osteoarthritis and type 2 diabetes; and five risk factors: alcohol consumption, BMI, cholesterol, smoking, systolic blood pressure (**Figure 1** and **Supplementary Table 1**).

**Table 1.**
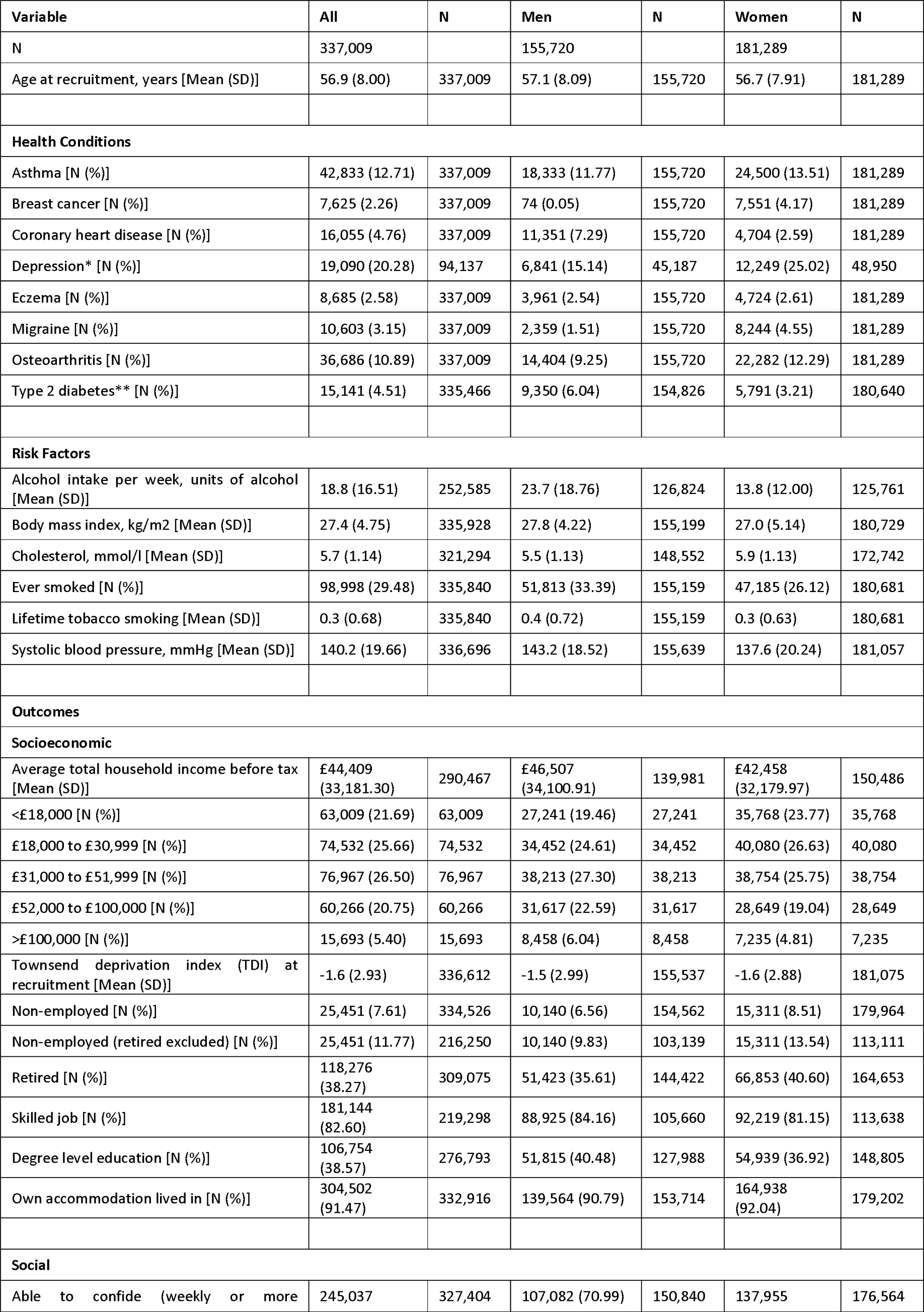
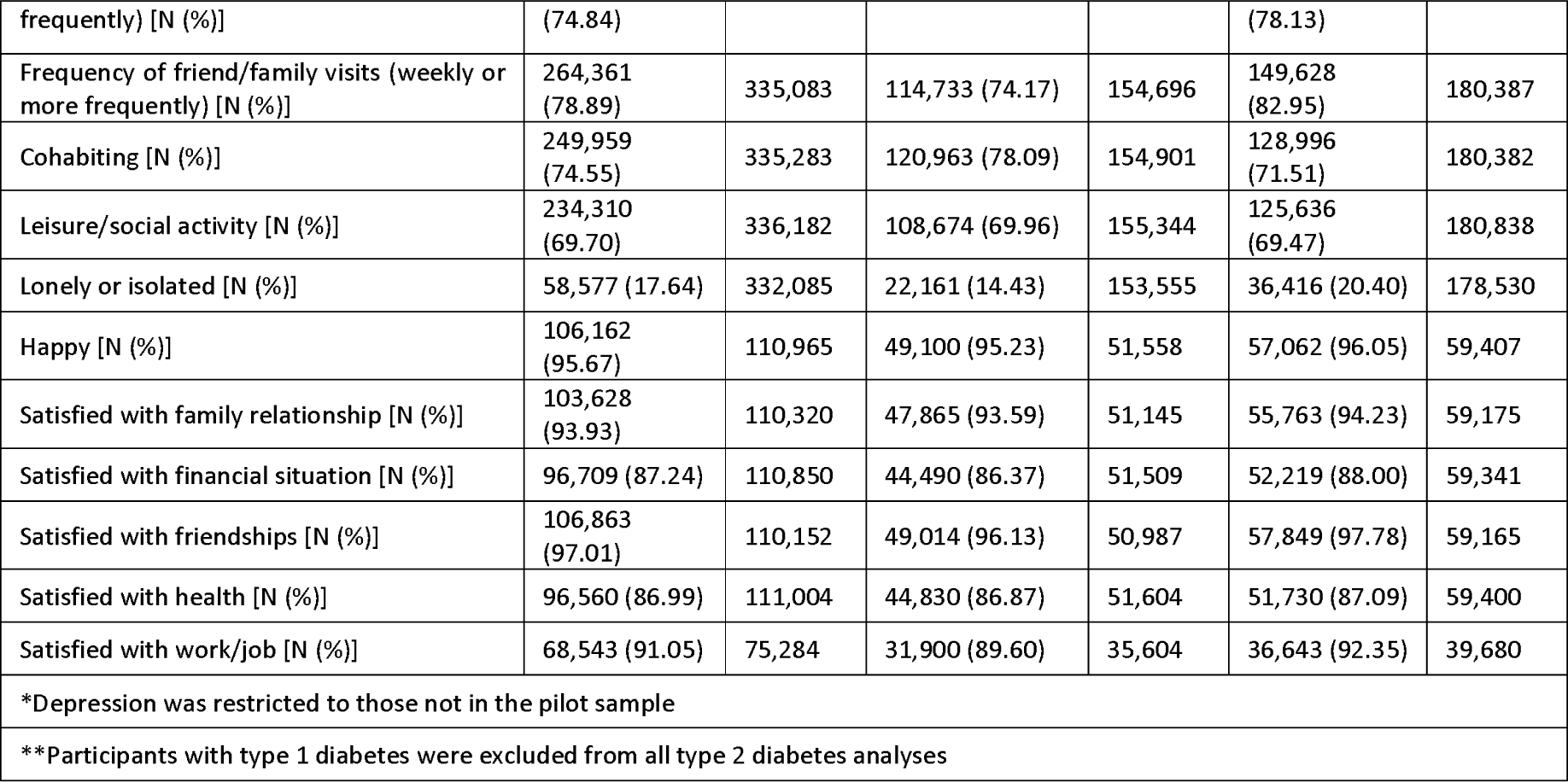
Summary demographics

**Figure 1.**
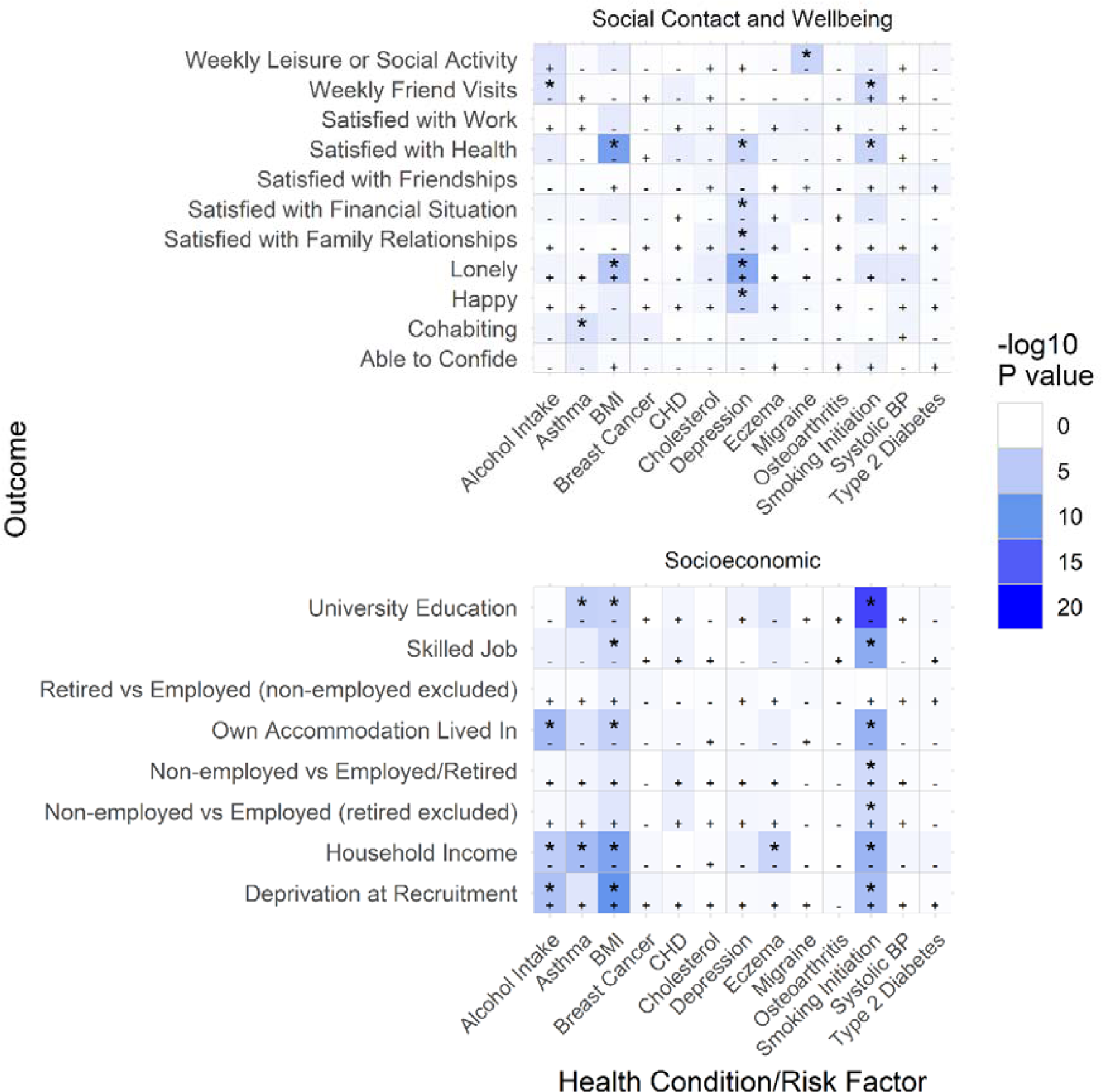
Heat map of results from the main analysis, smaller P values are shown by darker cells, starred results are below the Bonferroni-corrected P value threshold, negative effect directions are denoted with a “-” sign, and positive effect directions are denoted with a “+” sign

Except for depression, we categorised a participant as having a health condition if they reported the condition at the baseline visit, or if they had the corresponding HES or cancer registry ICD-9 or ICD-10 code for the health condition before the baseline visit (ICD codes and specific questions used shown in **Supplementary Table 1**).

We coded depression as in Tyrrell (17), where participants were considered to have depression if they self-reported seeing a GP or psychiatrist for nerves, anxiety or depression and reported at least a 2-week duration of depression or unenthusiasm, or had the relevant ICD-9 or ICD-10 codes for depression. Participants were considered to not have depression if they did not report ever visiting a GP or psychiatrist for nerves, anxiety or depression, did not self-report having depression and did not have an ICD code for depression. Only 10 centres asked the questions related to depression, so only participants from these centres were considered in the depression analyses.

The measurement of health risk factors is described in **Box 1**.

#### Box 1: Measurement of health risk factors at baseline (except smoking variables)

##### Alcohol intake

We estimated the average weekly intake of alcoholic units (10ml of pure alcohol) for all participants based on the average reported intake of six different types of alcoholic beverage. The nominal number of units we assigned per drink for each type of alcoholic beverage are listed below:

- Red wine: 125 ml (6/bottle), 14% = 1.75 units
- Champagne/white wine: 125 ml (6/bottle), 14% = 1.75 units
- Beer/cider: 1 pint, 3.5% = 2 units
- Spirits: 25 ml (25 standard measures in a normal sized bottle), 40% = 1 unit
- Fortified wine: 60 ml (12/bottle), 20% = 1.2 units
- Other: Unknown, example is an alcopop = 1 unit

We removed self-reported former drinkers, participants with a very high number of units per week (>200 units), and participants who did not report they were never drinkers but who answered none of the questions about weekly alcohol intake, leaving 252,585 participants (75%).

##### Body mass index

BMI was estimated as measured weight in kilograms divided by measured height in metres squared.

##### Cholesterol

Cholesterol was measured by UK Biobank at baseline (measured by CHO-POD analysis on a Beckman Coulter AU5800).

##### Smoking

We used two measures of self-reported smoking.

###### Lifetime smoking index

a composite (continuous) measure of relevant smoking variables with a simulated half-time constant representing the decreasing effect of smoking on health outcomes over time. This variable was created by Wootton and colleagues and used in a paper studying smoking and depression/schizophrenia (18).

###### Smoking initiation

a binary measure indicating whether participants had ever versus never smoked participants, based on whether the lifetime smoking index value had a non-zero value.

##### Systolic blood pressure

Systolic blood pressure was measured using an automated device, and two measurements were taken a few moments apart. If the standard automated device could not be employed, two manual readings were taken instead.

### Polygenic Risk Scores (Instrumental Variables)

We searched previous genome-wide association studies (GWAS) for single nucleotide polymorphisms (SNPs) with strong evidence of associations for each health condition and risk factor, defined as having a P value at genome-wide significance (P ≤ 5 × 10^−8^) (further details in **Supplementary Information 2** and **Supplementary Tables 2 and 3**). The polygenic risk scores (PRS) for each health condition and risk factor were then calculated as the sum of the effect alleles for all SNPs associated with the health condition or risk factor, with each SNP weighted by the regression coefficient from the GWAS from which the SNP was identified.

**Table 2.** Main MR, split-sample MR and multivariable adjusted analysis results for all outcomes where the main or split-sample Mendelian randomization analysis had a P value less than 0.0026

| Health Condition/Risk Factor | Outcome | N | Main MR Analysis |  | Split-Sample MR Analysis |  | Multivariable Adjusted Analysis |  | P for Endogeneity |
| --- | --- | --- | --- | --- | --- | --- | --- | --- | --- |
|  |  |  | Beta (95% CI) | P | Beta (95% CI) | P | Beta (95% CI) | P |  |
| Alcohol Intake (5 units/week) | Cohabiting | 251519 | -1.0% (-2.1% to 0.2%) | 9.21E-02 | -1.8% (-2.8% to -0.8%) | **3.16E-04** | -0.4% (-0.4% to -0.3%) | 0 | 2.98E-01 |
| Alcohol Intake (5 units/week) | Household Income | 219929 | £-2111 (£-3093 to £-1128) | **2.55E-05** | £ 413 (£-406 to £1233) | 3.23E-01 | £ 442 (£ 400 to £ 484) | 0 | **1.62E-07** |
| Alcohol Intake (5 units/week) | Own Accommodation Lived In | 249893 | -1.7% (-2.4% to -1.0%) | **5.98E-07** | -0.9% (-1.5% to -0.3%) | **1.89E-03** | -0.2% (-0.2% to -0.1%) | 4.29E-25 | **4.28E-06** |
| Alcohol Intake (5 units/week) | TDI at Recruitment | 252294 | 0.18 (0.10 to 0.25) | **1.92E-06** | 0.13 (0.07 to 0.19) | **4.97E-05** | 0.03 (0.03 to 0.04) | 0 | **8.05E-05** |
| Alcohol Intake (5 units/week) | Weekly Friend Visits | 251271 | -1.8% (-2.9% to -0.6%) | **2.34E-03** | -0.8% (-1.7% to 0.2%) | 1.19E-01 | -0.0% (-0.1% to 0.0%) | 8.95E-02 | **2.73E-03** |
| Asthma | Cohabiting | 335283 | -11.0% (-18.0% to -4.0%) | **2.00E-03** | -1.8% (-6.8% to 3.3%) | 4.93E-01 | -1.6% (-2.0% to -1.1%) | 1.68E-12 | **7.90E-03** |
| Asthma | Household Income | 290467 | £-13519 (£-18794 to £-8244) | **5.09E-07** | £-6690 (£-10506 to £-2874) | **5.90E-04** | £-694 (£-1030 to £-358) | 5.21E-05 | **1.44E-06** |
| Asthma | University Education | 276793 | -17.0% (-25.3% to -8.7%) | **5.93E-05** | -12.4% (-18.5% to -6.4%) | **5.49E-05** | 2.1% (1.6% to 2.7%) | 1.14E-14 | **4.96E-06** |
| Body Mass Index (5 kg/m<sup>2</sup>) | Household Income | 289604 | £-2778 (£-3693 to £-1864) | **2.60E-09** | £-4299 (£-5158 to £-3440) | **1.05E-22** | £-2332 (£-2451 to £-2213) | 0 | 3.35E-01 |
| Body Mass Index (5 kg/m<sup>2</sup>) | Lonely | 331030 | 2.4% (1.4% to 3.5%) | **7.61E-06** | 2.9% (1.9% to 3.9%) | **1.50E-08** | 2.7% (2.6% to 2.9%) | 0 | 5.43E-01 |
| Body Mass Index (5 kg/m<sup>2</sup>) | Non-employed vs Employed (retired excluded) | 215634 | 1.4% (0.4% to 2.5%) | 5.84E-03 | 2.7% (1.7% to 3.7%) | **6.56E-08** | 1.9% (1.8% to 2.0%) | 0 | 3.67E-01 |
| Body Mass Index (5 kg/m<sup>2</sup>) | Non-employed vs Employed/Retired | 333458 | 0.9% (0.2% to 1.6%) | 1.37E-02 | 1.8% (1.2% to 2.5%) | **1.57E-07** | 1.3% (1.2% to 1.4%) | 0 | 2.49E-01 |
| Body Mass Index (5 kg/m<sup>2</sup>) | Own Accommodation Lived In | 331866 | -1.6% (-2.4% to -0.8%) | **5.79E-05** | -3.3% (-4.0% to -2.6%) | **3.82E-19** | -2.6% (-2.7% to -2.5%) | 0 | **9.31E-03** |
| Body Mass Index (5 kg/m<sup>2</sup>) | Satisfied with Financial Situation | 110380 | -1.6% (-3.2% to 0.0%) | 5.48E-02 | -3.0% (-4.6% to -1.5%) | **1.19E-04** | -3.4% (-3.6% to -3.2%) | 0 | 2.55E-02 |
| Body Mass Index (5 kg/m<sup>2</sup>) | Satisfied with Health | 110520 | -5.1% (-6.8% to -3.5%) | **9.91E-10** | -6.7% (-8.3% to -5.1%) | **4.17E-17** | -7.6% (-7.8% to -7.4%) | 0 | **3.38E-03** |
| Body Mass Index (5 kg/m<sup>2</sup>) | Skilled Job | 218819 | -2.2% (-3.5% to -1.0%) | **4.69E-04** | -5.5% (-6.7% to -4.3%) | **5.61E-19** | -2.3% (-2.5% to -2.1%) | 0 | 9.32E-01 |
| Body Mass Index (5 kg/m<sup>2</sup>) | TDI at Recruitment | 335536 | 0.25 (0.18 to 0.33) | **7.73E-11** | 0.35 (0.28 to 0.42) | **1.08E-21** | 0.29 (0.28 to 0.30) | 0 | 2.98E-01 |
| Body Mass Index (5 kg/m<sup>2</sup>) | University Education | 276030 | -2.9% (-4.4% to -1.5%) | **9.82E-05** | -9.3% (-10.7% to -7.9%) | **6.42E-39** | -5.4% (-5.5% to -5.2%) | 0 | **1.10E-03** |
| Body Mass Index (5 kg/m<sup>2</sup>) | Weekly Leisure or Social Activity | 335108 | -1.4% (-2.7% to -0.1%) | 2.91E-02 | -4.6% (-5.8% to -3.4%) | **5.27E-14** | -2.7% (-2.8% to -2.5%) | 0 | 5.46E-02 |
| Depression | Happy | 69649 | -19.1% (-28.3% to -9.8%) | **5.32E-05** |  |  | -7.4% (-7.7% to -7.0%) | 0 | **9.83E-03** |
| Depression | Lonely | 92983 | 58.7% (38.6% to 78.9%) | **1.15E-08** |  |  | 18.4% (17.8% to 19.0%) | 0 | **1.88E-05** |
| Depression | Satisfied with Family Relationships | 69275 | -19.2% (-30.4% to -8.1%) | **7.44E-04** |  |  | -6.9% (-7.2% to -6.5%) | 0 | 2.57E-02 |
| Depression | Satisfied with Financial Situation | 69557 | -26.5% (-41.9% to -11.0%) | **7.88E-04** |  |  | -9.1% (-9.7% to -8.6%) | 0 | 2.39E-02 |
| Depression | Satisfied with Health | 69653 | -29.2% (-44.7% to -13.7%) | **2.24E-04** |  |  | -11.1% (-11.6% to -10.5%) | 0 | 1.82E-02 |
| Eczema | Household Income | 290467 | £-46987 (£-71048 to £-22925) | **1.29E-04** | £-6774 (£-25198 to £11649) | 4.71E-01 | £ 157 (£-544 to £ 859) | 6.60E-01 | **7.36E-05** |
| Lifetime Smoking (SD) | Cohabiting | 334133 |  |  | -5.3% (-8.7% to -1.9%) | **2.01E-03** | -4.5% (-4.7% to -4.4%) | 0 |  |
| Lifetime Smoking (SD) | Household Income | 289737 |  |  | £-7739 (£-10308 to £-5170) | **3.53E-09** | £-4089 (£-4201 to £-3976) | 0 |  |
| Lifetime Smoking (SD) | Non-employed vs Employed (retired excluded) | 215673 |  |  | 5.8% (2.8% to 8.9%) | **1.83E-04** | 4.1% (4.0% to 4.2%) | 0 |  |
| Lifetime Smoking (SD) | Non-employed vs Employed/Retired | 333395 |  |  | 4.1% (2.0% to 6.2%) | **1.22E-04** | 2.7% (2.6% to 2.8%) | 0 |  |
| Lifetime Smoking (SD) | Own Accommodation Lived In | 331787 |  |  | -8.7% (-10.9% to -6.6%) | **2.87E-15** | -5.1% (-5.2% to -5.0%) | 0 |  |
| Lifetime Smoking (SD) | Satisfied with Financial Situation | 110508 |  |  | -9.5% (-14.3% to -4.8%) | **8.39E-05** | -4.0% (-4.2% to -3.8%) | 0 |  |
| Lifetime Smoking (SD) | Satisfied with Health | 110656 |  |  | -8.3% (-13.2% to -3.4%) | **9.46E-04** | -3.7% (-3.9% to -3.5%) | 0 |  |
| Lifetime Smoking (SD) | Skilled Job | 218702 |  |  | -9.0% (-13.1% to -4.8%) | **2.13E-05** | -4.0% (-4.2% to -3.9%) | 0 |  |
| Lifetime Smoking (SD) | TDI at Recruitment | 335445 |  |  | 0.99 (0.77 to 1.20) | **1.58E-19** | 0.53 (0.52 to 0.54) | 0 |  |
| Lifetime Smoking (SD) | University Education | 276185 |  |  | -16.2% (-20.9% to -11.4%) | **2.68E-11** | -6.0% (-6.2% to -5.8%) | 0 |  |
| Migraine | Weekly Leisure or Social Activity | 336182 | -43.7% (-66.0% to -21.3%) | **1.31E-04** | -26.3% (-57.7% to 5.1%) | 1.01E-01 | -2.9% (-3.8% to -2.0%) | 2.04E-10 | **2.98E-04** |
| Smoking Initiation | Household Income | 289737 | £-24394 (£-33403 to £-15384) | **1.11E-07** | £-19188 (£-26182 to £-12195) | **7.54E-08** | £-5974 (£-6220 to £-5728) | 0 | **3.21E-05** |
| Smoking Initiation | Non-employed vs Employed (retired excluded) | 215673 | 19.2% (8.5% to 29.9%) | **4.20E-04** | 8.6% (0.4% to 16.9%) | 3.95E-02 | 5.4% (5.0% to 5.7%) | 0 | **9.49E-03** |
| Smoking Initiation | Non-employed vs Employed/Retired | 333395 | 13.0% (5.7% to 20.3%) | **4.85E-04** | 5.7% (0.2% to 11.2%) | 4.26E-02 | 3.6% (3.4% to 3.7%) | 0 | 1.01E-02 |
| Smoking Initiation | Own Accommodation Lived In | 331787 | -21.5% (-29.3% to -13.6%) | **8.10E-08** | -16.8% (-22.5% to -11.0%) | **1.05E-08** | -6.7% (-6.9% to -6.5%) | 0 | **1.39E-04** |
| Smoking Initiation | Satisfied with Financial Situation | 110508 | -21.2% (-36.9% to -5.4%) | 8.33E-03 | -22.5% (-36.2% to -8.8%) | **1.29E-03** | -5.7% (-6.2% to -5.3%) | 0 | 4.90E-02 |
| Smoking Initiation | Satisfied with Health | 110656 | -32.4% (-48.9% to -15.8%) | **1.28E-04** | -22.0% (-35.9% to -8.0%) | **2.04E-03** | -5.6% (-6.0% to -5.2%) | 0 | **7.51E-04** |
| Smoking Initiation | Skilled Job | 218702 | -40.5% (-54.5% to -26.5%) | **1.50E-08** | -13.4% (-23.5% to -3.4%) | 8.67E-03 | -5.5% (-5.9% to -5.2%) | 0 | **1.26E-07** |
| Smoking Initiation | TDI at Recruitment | 335445 | 1.89 (1.13 to 2.64) | **9.61E-07** | 2.23 (1.66 to 2.81) | **2.76E-14** | 0.79 (0.77 to 0.81) | 0 | **3.91E-03** |
| Smoking Initiation | University Education | 276185 | -73.8% (-90.7% to -56.9%) | **1.22E-17** | -37.3% (-48.7% to -26.0%) | **1.26E-10** | -9.7% (-10.1% to -9.3%) | 0 | **5.00E-18** |
| Smoking Initiation | Weekly Friend Visits | 333966 | 21.1% (9.7% to 32.5%) | **2.76E-04** | -1.9% (-10.2% to 6.4%) | 6.49E-01 | -0.4% (-0.7% to -0.1%) | 4.39E-03 | **1.32E-04** |

### Covariates

Age, sex and UK Biobank recruitment centre were reported at the baseline assessment, and genetic principal components (used to control for population stratification (19)) were derived by UK Biobank.

### Social and Socioeconomic Measures (Outcomes)

We selected social and socioeconomic outcomes measured at the UK Biobank baseline assessment centre. Where possible, we dichotomised outcomes to simplify interpretability and comparability across outcomes. **Box 2** contains a list of all outcomes; **Supplementary Information 3** and **Supplementary Table 4** give further information on how each outcome was measured.

We considered breast cancer, coronary heart disease, osteoarthritis, cholesterol or systolic blood pressure unlikely to have plausible causal effects on the chance of obtaining a university degree given that these health conditions usually occur later in life; the Mendelian randomization effect estimates for these associations were thus used as negative controls (i.e. where no effect should be expected) (20,21).

#### Box 2: List of all social and socioeconomic measures (outcomes)

##### Socioeconomic Outcomes

- Average household income before tax, with each category assigned the mid-point of the range (and open-ended categories a nominal value) to allow for continuous analysis*:
  ∘ <£18,000 = £15,000
  ∘ 18,000 to £30,999 = £24,500
  ∘ 31,000 to £51,999 = £41,500
  ∘ 52,000 to £100,000 = £76,000
  ∘ >£100,000 = £150,000
- Deprivation, measured using the Townsend Deprivation Index (TDI) of current address*
- Current employment status, coded as three separate outcomes:
  ∘ Non-employed, not retired (versus employed or retired)
  ∘ Non-employed (versus employed, retired excluded)
  ∘ Retired (versus still employed, other non-employed excluded)
- Job class, coded as skilled versus unskilled (22)
- Degree status, coded as degree-level education versus lower
- Owner-occupied accommodation versus renting

##### Social Outcomes

###### Measures of social contact

- Having someone to confide in weekly or more frequently versus less frequently
- Friend/family visits weekly or more frequently versus less frequently
- Cohabiting with partner or spouse versus not cohabiting
- Participation in any leisure/social activity versus none

###### Measures of happiness and wellbeing

- Lonely/isolated versus not lonely/isolated
- Extremely/very/moderately happy versus not
- Extremely/very/moderately happy with family relationship versus not
- Extremely/very/moderately happy with financial situation versus not
- Extremely/very/moderately happy with friendships versus not
- Extremely/very/moderately happy with health versus not
- Extremely/very/moderately happy with work/job versus not

*Income and deprivation were both dichotomised as additional analyses so the results could be included in plots of all results comparing across outcomes: ≥£52,000 versus <£52,000 for income, most deprived third of TDI versus two least deprived thirds for deprivation

### Main Mendelian Randomization Analysis

We used Mendelian randomization to estimate the causal association between each health condition and risk factor and each outcome, using the PRS as an instrumental variable, with age at baseline assessment, sex, UK Biobank recruitment centre and 40 genetic principal components as covariates. We used the ivreg2 package in Stata (version 15.1) with robust standard errors, and tested for weak instrument bias (using Kleibergen-Paap Wald rk F statistics) to assess whether the PRS were sufficiently predictive of the exposures (23). This Mendelian randomization analysis estimates mean and risk differences for continuous and binary outcomes respectively using additive structural mean models (24–26). Mean differences are interpreted as the average change in the outcome over all participants for having the exposure, and risk differences are interpreted as the absolute percentage point change in proportion of participants with the outcome for having the exposure (as in a linear probability model). For health conditions, we are measuring the effects of genetic liability to the health condition (27). The analysis of breast cancer as an exposure was restricted to women. Despite the limitations of an approach based on statistical significance (28), the number of results generated in these analyses necessitated a decision about which results to present in the main paper. Therefore, in the main table of results, we report results with a P value less than 0.0026 (a Bonferroni-corrected P value of 0.05 divided by 19 outcomes, with no correction for multiple exposures), while full results are reported in supplementary tables. However, we considered the public health implications of all effect estimates when interpreting results.

To compare the Mendelian randomization results with associations from non-genetic analysis, we estimated the multivariable adjusted associations between the exposures and outcomes using linear regression, with age, sex, recruitment centre and 40 genetic principle components as covariates, i.e. observational analyses without genetic variables. These are linear probability models for binary outcomes (rather than logistic regression models), which were necessary to be able to compare with the Mendelian randomization analyses, as they are equivalent to additive structural mean models. We also performed endogeneity tests (29) to test whether the Mendelian randomization and multivariable adjusted association estimates differed, where a low P value indicates there was evidence the Mendelian randomization and multivariable effects were different.

### Sensitivity Analyses

The robustness of Mendelian randomization analyses is reliant on the assumption that the SNPs, and therefore PRS, do not affect the outcome except through the exposure, i.e. the SNPs are not pleiotropic. We tested this assumption by conducting sensitivity Mendelian randomization analyses, including inverse-variance weighted (IVW), MR Egger (an indicator of directional pleiotropy), weighted median, weighted mode and simple mode analyses (30–32). We also measured Cochran’s Q statistic from the IVW analyses (a measure of heterogeneity in the effects of individual SNPs on the outcome), an indicator of pleiotropy (33) or problems with modelling assumptions (34).

From these analyses, we determined: a) whether the results were consistent with the main Mendelian randomization analysis, which would indicate the results of the main analysis were robust, and b) whether there was evidence of pleiotropy from both the Egger regression constant term and Cochran’s Q statistic. We also visually inspected plots of the sensitivity Mendelian randomization analyses, which would indicate possible bias in the results of the main analysis. Sensitivity Mendelian randomization analyses could only be performed when there were three or more SNPs included in each PRS.

We also conducted split-sample GWAS and Mendelian randomization analysis using UK Biobank data, in which we randomly split UK Biobank into halves, and for each half conducted a GWAS for each health condition and risk factor using the MRC IEU UK Biobank GWAS pipeline (35). The results of the two GWAS were used to create PRS for the other half of UK Biobank avoiding sample overlap (36), and we repeated the Mendelian randomization analysis with the two PRS separately, then combined the two results with fixed-effect meta-analysis to give a single estimate. The split-sample analysis a) allowed us to analyse lifetime smoking, as this has only been generated in UK Biobank, and thus no previous GWAS could have been used to inform the PRS, b) allowed us to potentially increase the size and power of the GWAS, possibly improving the predictive ability of the PRS and c) guaranteed homogeneity of the GWAS and analysis populations, which removes the potential bias from using data from an external GWAS to inform the creation of the PRS, for example, through differences in populations giving different effects of SNPs. We also performed sensitivity Mendelian randomization sensitivity analyses on each split to check the robustness of the split-sample results.

**Supplementary Table 5** shows a summary of all PRS created and used in the split-sample analyses, and all GWAS significant SNPs from the split-sample GWAS are detailed in **Supplementary Table 6**.

### Secondary Analyses

We conducted secondary analyses to check the robustness of results, looking at whether: a) results are different by sex and deprivation at birth, b) results for household income are affected by household size (income equivalisation), c) results for employment outcomes are different when restricting to working age participants, d) results for household income are different when restricting to participants who have not retired and e) results for smoking are robust when only looking at the SNP rs1051730, known to affect smoking heaviness (37). Further information for the secondary analyses and results are in **Supplementary Information 4**.

### Patient and Public Involvement

This study was conducted using UK Biobank. Details of patient and public involvement in the UK Biobank are available online (www.ukbiobank.ac.uk/about-biobank-uk/ and https://www.ukbiobank.ac.uk/wp-content/uploads/2011/07/Summary-EGF-consultation.pdf?phpMyAdmin=trmKQlYdjjnQIgJ%2CfAzikMhEnx6). No patients were specifically involved in setting the research question or the outcome measures, nor were they involved in developing plans for recruitment, design, or implementation of this study. No patients were asked to advise on interpretation or writing up of results. There are no specific plans to disseminate the results of the research to study participants, but the UK Biobank disseminates key findings from projects on its website.

### Data and Code Availability

The empirical dataset will be archived with UK Biobank and made available to individuals who obtain the necessary permissions from the study’s data access committees. The code used to clean and analyse the data is available here: https://github.com/sean-harrison-bristol/Effects-of-Health-Conditions-and-Risk-Factors-on-Socioeconomic-Outcomes

## 3. Results

Summary demographics, including prevalence of health conditions, risk factors and all outcomes, are presented in **Table 1**. The mean age of participants was 56.6 years (standard deviation: 8.0 years), mean household income (estimated from household income category midpoints) was £44,409 (standard deviation: £33,181,), and 46% of participants were male. Results from the main Mendelian randomization analysis are displayed in a heat map of the P values, **Figure 1. Table 2** shows results from the main Mendelian randomization, split-sample Mendelian randomization and multivariable adjusted analyses for all outcomes where the main or split-sample Mendelian randomization analysis had a P value less than 0.0026. All health conditions (except osteoarthritis) and risk factors in the main Mendelian randomization analysis had a low risk of weak instrument bias, and 75% of regressions had F-statistics above 1000.

Forest plots showing the results for the main Mendelian randomization, split-sample Mendelian randomization and multivariable adjusted analyses for health conditions and risk factors on household income are shown in **Figures 2 and 3**, although there was evidence of heterogeneity between SNPs in sensitivity Mendelian randomization analyses for all exposures on income (Cochran’s Q statistic P < 0.0001), indicating possible pleiotropy. As such, results for income should be interpreted with some caution, especially if there is evidence of directional pleiotropy from MR Egger analyses. As additional examples, the main Mendelian randomization, split-sample Mendelian randomization and multivariable adjusted analyses for health conditions and risk factors on loneliness are shown in **Figures 4 and 5**; plots for all other analyses are presented in **Supplementary Materials**.

**Figure 2.**
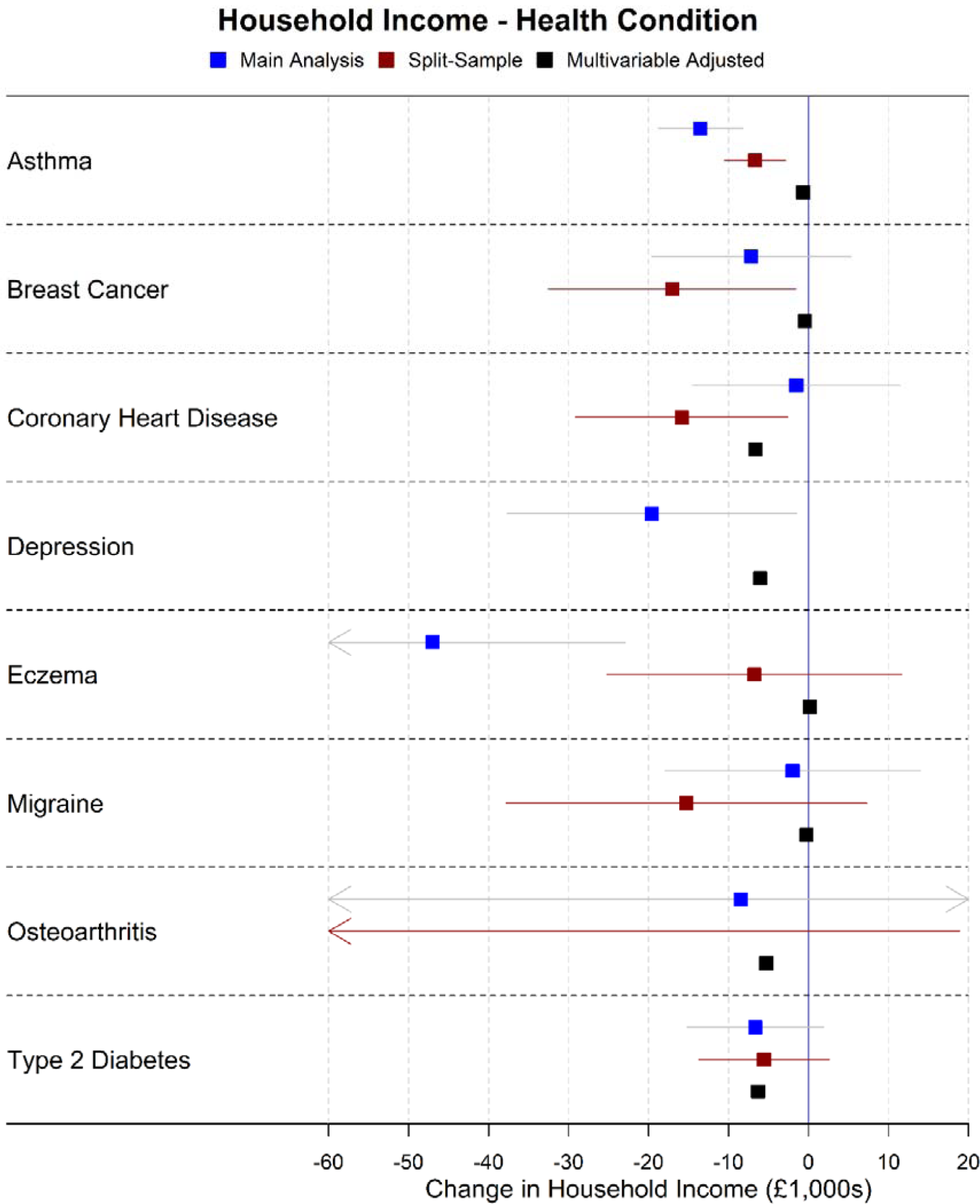
Forest plot showing effects of health conditions on household income for the main Mendelian randomization, split-sample Mendelian randomization and multivariable adjusted analyses (note: confidence intervals are so narrow for the multivariable adjusted analyses they cannot be seen)

**Figure 3.**
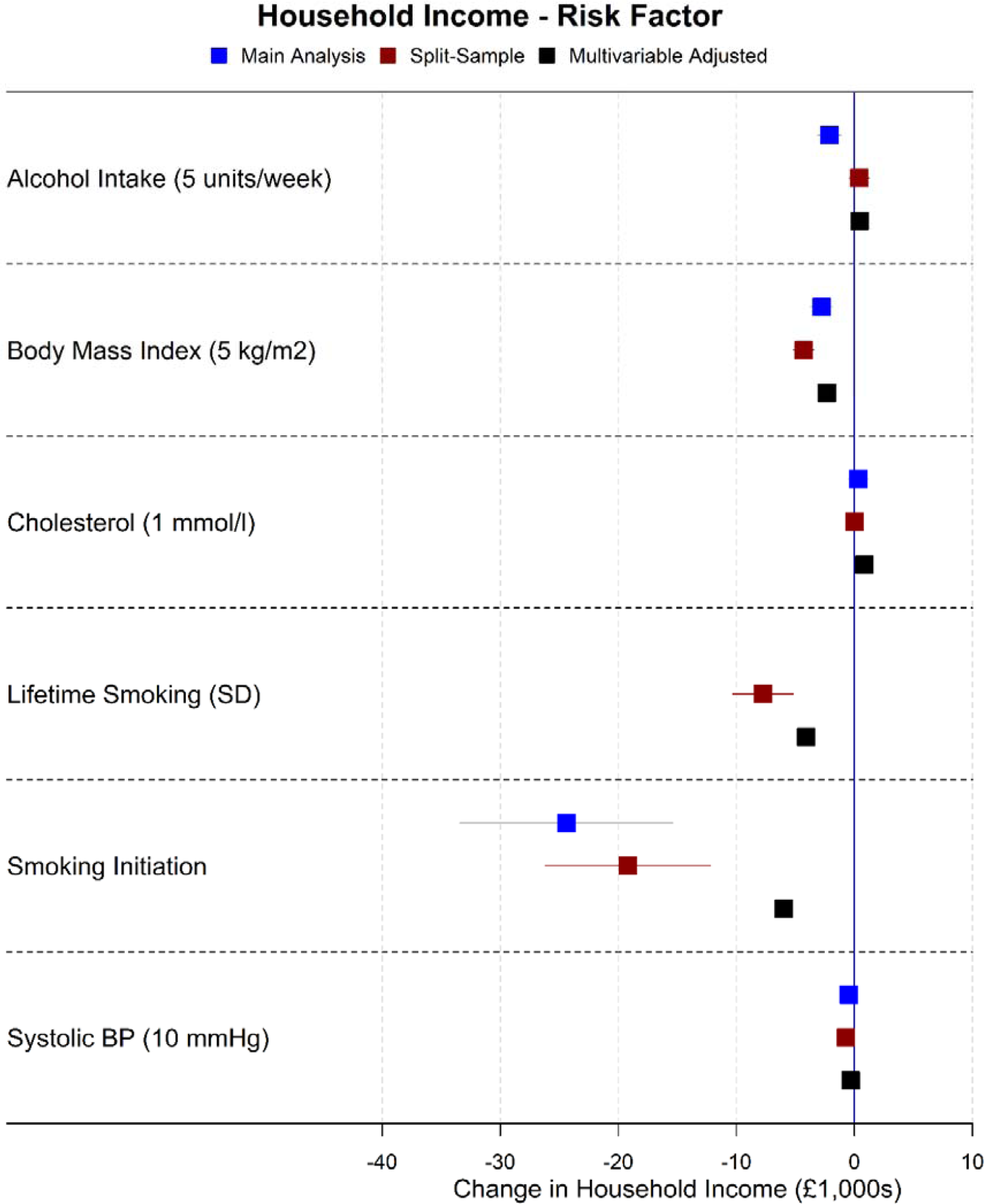
Forest plot showing effects of risk factors on household income for the main Mendelian randomization, split-sample Mendelian randomization and multivariable adjusted analyses (note: confidence intervals are so narrow they cannot be seen for most associations)

**Figure 4.**
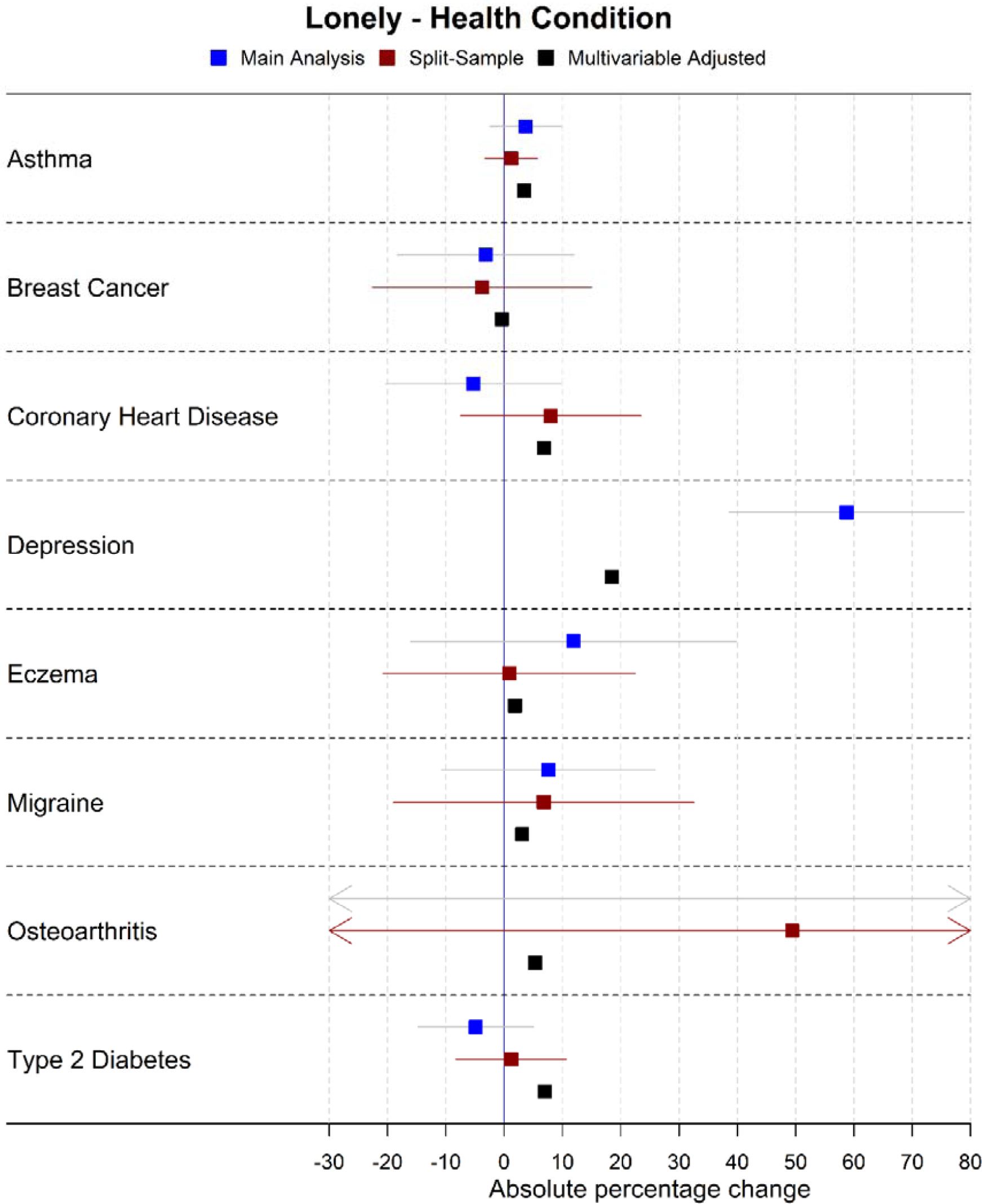
Forest plot showing effects of health conditions on being lonely for the main Mendelian randomization, split-sample Mendelian randomization and multivariable adjusted analyses (note: confidence intervals are so narrow for the multivariable adjusted analyses they cannot be seen)

**Figure 5.**
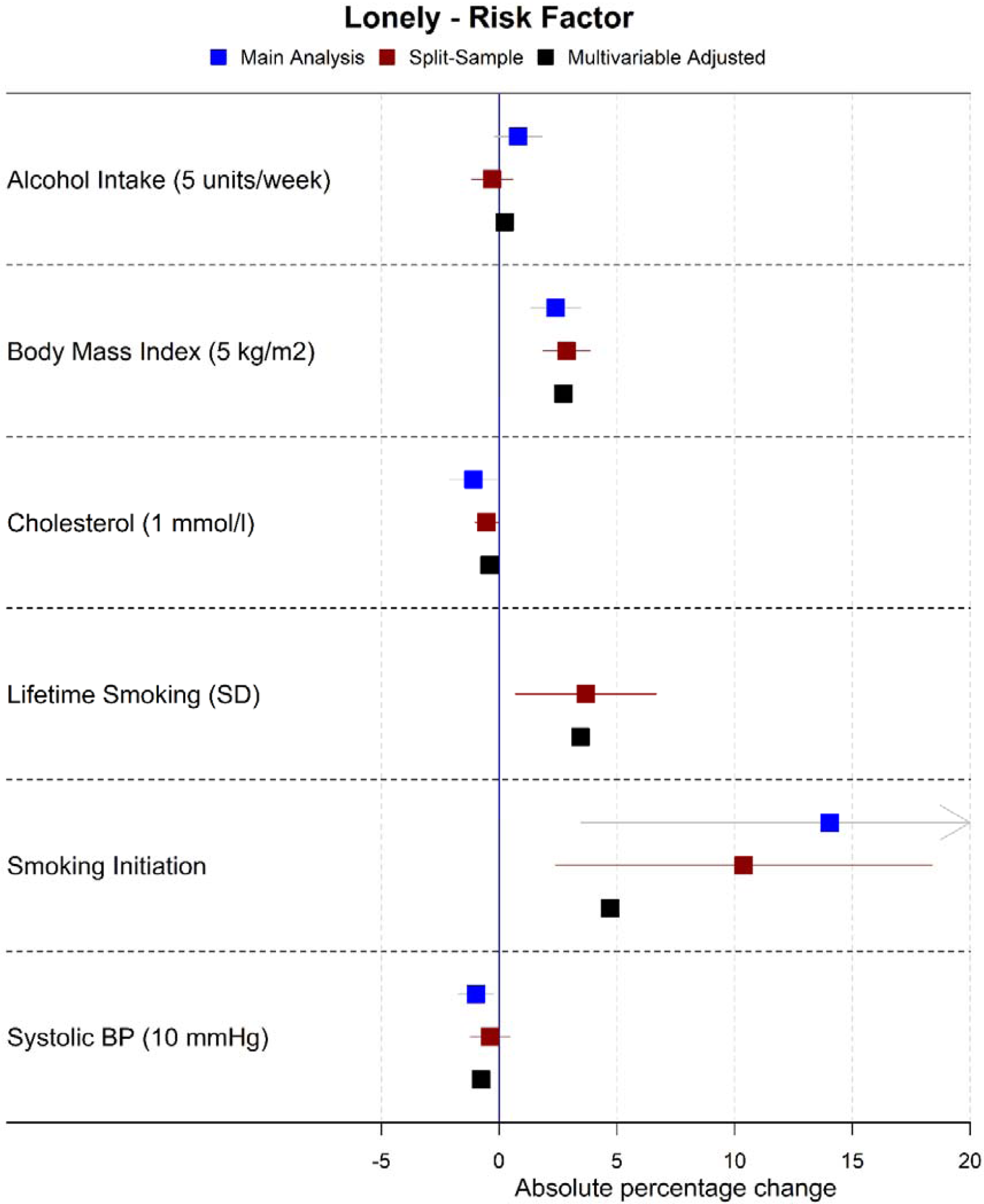
Forest plot showing effects of risk factors on being lonely for the main Mendelian randomization, split-sample Mendelian randomization and multivariable adjusted analyses (note: confidence intervals are so narrow for the multivariable adjusted analyses they cannot be seen)

### 3.1 Health Conditions

#### Asthma

In the main Mendelian randomization analysis, asthma was estimated to reduce household income (mean difference = −£13,519, 95% confidence interval (CI): −£18,794 to −£8,243), the chance of obtaining a university degree (absolute percentage change [APC] = −17.0%, 95% CI: −25.3% to −8.7%), and the chance of cohabiting (APC = −11.0%, 95% CI: −18.0% to −4.0%). There was little evidence asthma affected other outcomes. Split-sample Mendelian randomization analysis estimates similarly showed detrimental estimates of asthma on obtaining a university degree and income, but not on cohabiting, and there was only evidence of pleiotropy in sensitivity Mendelian randomization analyses for income. The multivariable adjusted association estimates tended to be weaker than the Mendelian randomization estimates, and in some cases (e.g. the chance of obtaining a university degree) in the opposite direction.

#### Depression

In the main Mendelian randomization analysis, depression was estimated to reduce satisfaction with health (APC = −29.2%, −44.7% to −13.7%), financial situation (APC = −26.5%, 95% CI: −41.9% to −11.0%) and family relationships (APC = −19.2%, 95% CI: −30.4% to −8.1%), and, as expected, reduce the chance of being happy (APC = −19.1%, 95% CI: −28.3% to −9.8%) and increase the chance of being lonely (APC = 58.7%, 95% CI: 38.6% to 78.9%). CIs were wide, but the point estimates were consistent with depression being detrimental for almost all socioeconomic outcomes, including household income (mean difference = −£19,584, 95% CI: −£37,679 to −£1,489). Depression was excluded from the split-sample analyses as no GWAS-significant SNPs were found in either split. There was evidence of heterogeneity in SNP effects for all outcomes, but no evidence of directional pleiotropy from Egger regression. Multivariable adjusted association estimates tended to be weaker than Mendelian randomization estimates.

#### Eczema

In the main Mendelian randomization analysis, eczema was estimated to reduce household income (mean difference = −£46,987, 95% CI: −£71,048 to −£22,925). However, this was not observed in the split-sample Mendelian randomization analysis (mean difference = £-6,774, 95% CI: £-25,198 to £11,649) or multivariable adjusted analysis (mean difference = £157, 95% CI: £-544 to £859). CIs for all other outcomes were very wide.

#### Migraine

In the main Mendelian randomization analysis, migraines were estimated to reduce the chance of having a weekly leisure or social activity (APC = −43.7%, 95% CI: −66.0% to −21.3%). This estimate was smaller in the split-sample Mendelian randomization (APC = −26.3% 95% CI: −57.7% to 5.1%) and multivariable regression analyses (APC = −2.9%, 95% CI: −3.8% to −2.0%). The CIs in Mendelian randomization analyses were wide for all other outcomes. There was no evidence of pleiotropy.

#### Type 2 Diabetes

In the main and split-sample Mendelian randomization analyses, there were no strong associations for type 2 diabetes with any outcome. Directions of effects were inconsistent across outcomes. Multivariable adjusted association estimates tended to be larger than Mendelian randomization estimates, and associations were apparent with several outcomes, most notably satisfaction with health (APC for multivariable adjusted association estimate = −19.1%, 95% CI: −20.1% to −18.2%).

#### Other Health Conditions

The CIs in Mendelian randomization analyses for breast cancer, coronary heart disease and osteoarthritis were very wide for all outcomes, and as such, these analyses were inconclusive. For breast cancer and coronary heart disease, there was no clear pattern of the direction of effects across outcomes, and CIs were wide. The CIs for osteoarthritis were very wide for all outcomes. As expected, given life course temporal relationships, there was little evidence from the main or split-sample Mendelian randomization analyses that breast cancer, coronary heart disease or osteoarthritis were associated with the chance of obtaining a university degree (included as negative controls). In the multivariable adjusted analysis, breast cancer was not associated with the chance of obtaining a university degree, while coronary heart disease and osteoarthritis were (APC = −8.1%, 95% CI: −9.0% to −7.1% and APC = −6.3%, 95% CI: −6.9% to −5.6%, respectively), indicating, together with the null estimates from the Mendelian randomization analyses, possible social causation of the health conditions, rather than vice versa. Osteoarthritis was excluded from the sensitivity Mendelian randomization analysis as there were fewer than 3 GWAS-significant SNPs in the osteoarthritis GWAS.

In the multivariable adjusted analysis, breast cancer was only associated with increased chances of being non-employed and retired and a decreased satisfaction with health, whereas coronary heart disease and osteoarthritis were negatively associated with all economic outcomes and most social outcomes, though not satisfaction with friendships or work nor with weekly friend visits.

### 3.2 Risk Factors

#### Alcohol Intake

All results are expressed for a 5 units per week increase in alcohol intake.

In the main Mendelian randomization analysis, alcohol was estimated to reduce household income (mean difference = −£2,111, 95% CI: −£3,093 to −£1,128) and the chance of owning accommodation (APC = −1.7%, −2.4% to −1.0%), and increase deprivation (mean difference in TDI = 0.18, 95% CI: 0.10 to 0.25, approximately 23% of a decile of TDI). In the split-sample Mendelian randomization analysis, alcohol was estimated to reduce the chance of cohabiting (APC = −1.8%, 95% CI: −2.8% to −0.8%) and owning accommodation (APC = −0.9%, 95% CI: −1.5% to −0.3%) and increase deprivation (mean difference in TDI = 0.13, 95% CI: 0.07 to 0.19, approximately 16% of a decile of TDI). There was no evidence of causal effects on other outcomes. There was evidence of heterogeneity in SNP effects for some outcomes (not deprivation), but no evidence of directional pleiotropy in Egger regression. The multivariable adjusted analysis estimated that alcohol increased (rather than reduced) household income (mean difference = £442, 95% CI: £400 to £484, P value from endogeneity test = 1.6×10^−7^), and no associations were seen with other outcomes.

#### Body Mass Index

All results are expressed for a 5 kg/m^2^ increase in BMI.

In the main Mendelian randomization analysis, BMI was estimated to be detrimental for all socioeconomic outcomes: BMI was estimated to reduce household income (mean difference = −£2,778, 95% CI: −£3,693 to −£1,864), and the chance of owning accommodation (APC = −1.6%, 95% CI:-2.4% to −0.8%), being satisfied with health (APC = −5.1%, −6.8% to −3.5%), obtaining a university degree (APC = −2.9%, 95% CI: −4.4% to −1.5%), and having a skilled job (APC = −2.2%, 95% CI: −3.5% to −1.0%), and increase deprivation (mean difference in TDI = 0.25, 95% CI: 0.18 to 0.33, approximately 31% of a decile of TDI) and the chance of being lonely (APC = 2.4%, 95% CI: 1.4% to 3.5%). In the split-sample analysis, effects of BMI were estimated to be more detrimental than in the main analysis for the above associations, and additionally to increase the chance of being non-employed, both when including and excluding retired participants (APC = 1.8%, 95% CI: 1.2% to 2.5% and APC = 2.7%, 95% CI: 1.7% to 3.7%, respectively), and reduce the chance of being satisfied with financial situation (APC = −3.0%, 95% CI: −4.6% to −1.5%) and having a weekly leisure or social activity (APC = −4.6%, 95% CI: −5.8% to −3.4%).

There was evidence of heterogeneity in SNPs for all outcomes, but evidence of directional pleiotropy in Egger regression only for obtaining a university degree. The multivariable adjusted associations between BMI and socioeconomic outcomes were generally consistent with the Mendelian randomization estimates.

#### Cholesterol

All results are expressed for a 1 mmol/litre increase in cholesterol.

In the main and split-sample Mendelian randomization analyses, there was no evidence of effects of cholesterol on any outcome. In the multivariable adjusted analyses, cholesterol was beneficial for all socioeconomic outcomes and most social contact and wellbeing outcomes, which, together with the null estimates from the Mendelian randomization analyses, confounding or reverse causation in the multivariable adjusted association estimates.

#### Lifetime Smoking

All results are expressed for a one standard deviation increase in the continuous lifetime smoking index value. We did not perform a main Mendelian randomization analysis, as there was no previous GWAS for lifetime smoking.

In the split-sample Mendelian randomization analysis, smoking was estimated to reduce household income (mean difference = −£7,739, 95% CI: −£10,308 to −£5,170), the chance of cohabiting (APC = −5.3%, 95% CI: −8.7% to −1.9%), owning accommodation (APC = −8.7%, 95% CI: −10.9% to −6.6%), having a skilled job (APC = −9.0%, 95% CI: −13.1% to −4.8%), obtaining a university degree (APC = −16.2%, 95% CI: −20.9% to −11.4%), and being satisfied with one’s financial situation (APC = −9.5%, 95% CI: −14.3% to −4.8%) and health (APC = −8.3%, 95% CI: −13.2% to −3.4%). Lifetime smoking was also estimate to increase deprivation (mean difference in TDI = 0.99, 95% CI: 0.77 to 1.20, approximately 124% of a decile of TDI) and the chance of being non-employed, both with retired participants included and excluded (APC = 4.1%, 95% CI: 2.0% to 6.2% and APC = 5.8%, 95% CI: 2.8% to 8.9% respectively). There was little evidence smoking affected other social outcomes. There was evidence of heterogeneity in SNPs for all outcomes, but evidence of directional pleiotropy in Egger regression only for being satisfied with health and having a skilled job. Multivariable adjusted analyses showed smaller estimates for all outcomes.

#### Smoking Initiation

In the main Mendelian randomization analysis, smoking initiation was estimated to reduce household income (mean difference = −£24,394, 95% CI: −£33,403 to −£15,384), the chance of owning accommodation (APC = −21.5%, 95% CI: −29.3% to −13.6%), being satisfied with health (APC = −32.4%, 95% CI: −48.9% to −15.8%), and of obtaining a university degree (APC = −73.8%, 95% CI: −90.7% to −56.9%), and to increase deprivation (mean difference in TDI = 1.89, 95% CI: 1.13 to 2.64, approximately 236% of a decile of TDI). All effects were also seen in the split-sample analysis. Smoking initiation was also estimated to increase the chance of having a skilled job (APC = −40.5%, 95% CI: −54.5% to −26.5%), and reduce the chance of being non-employed, both including and excluding retired participants (APC = 13.0%, 95% CI: 5.7% to 20.3% and APC = 19.2%, 95% CI: 8.5% to 29.9% respectively), and of having weekly friend visits (APC = 21.1%, 95% CI: 9.7% to 32.5%), but only in the main Mendelian randomization analysis. Additionally, smoking initiation was estimated to reduce the chance of being satisfied with one’s financial situation (APC = −22.5%, 95% CI: −36.2% to −8.8%) in the split-sample Mendelian randomization analysis, with a similar effect size in the main Mendelian randomization analysis. CIs were wide for all outcomes. There was evidence of heterogeneity in SNP effects for all outcomes, but no evidence of directional pleiotropy from Egger regression. Multivariable adjusted association estimates tended to be closer to the null than the MR analyses.

#### Systolic BP

All results are expressed for a 10-mmHg increase in systolic blood pressure.

In the main and split-sample Mendelian randomization analyses, there was no evidence of effects of systolic BP on any outcome.

### 3.3 Further Analyses

Full results from main Mendelian randomization, sensitivity Mendelian randomization, split-sample Mendelian randomization, and split-sample sensitivity Mendelian randomization analyses are shown in **Supplementary Tables 7-10**. For all health conditions and risk factors, forest plots showing results for the main Mendelian randomization, split-sample Mendelian randomization and multivariable adjusted analyses (presented both as each exposure on social and socioeconomic outcomes, and for each outcome on health conditions and risk factors) are available in **Supplementary Materials**, along with forest plots of SNPs and plots showing IVW, MR Egger, simple mode, weighted median and weighted mode Mendelian randomization analyses.

## 4. Discussion

We estimate the putative causal effects of a variety of health conditions and risk factors on socioeconomic and social outcomes using Mendelian randomization, a genetically-informed methodology typically less affected by confounding and reverse causality than observational analyses that adjust for measured confounders (11). Our results indicate that higher BMI, greater alcohol intake and smoking all negatively affect socioeconomic outcomes, and depression negatively affects many social outcomes. We do not observe an effect of cholesterol or systolic BP on any outcome, which may reflect effective treatments for high cholesterol and hypertension protecting participants from adverse consequences. For breast cancer, coronary heart disease, migraine and osteoarthritis, the confidence intervals for all Mendelian randomization analyses were all very wide, meaning that it is not possible to draw firm conclusions about the social and socioeconomic consequences of these conditions from our analyses.

Potential reasons for adverse effects of high BMI, alcohol use and smoking on social and socioeconomic outcomes include increased disease burden, social stigma (e.g. bias against obese people, smokers etc.), or behaviours which make employment, retention of employment, or social interaction challenging. Our previous analyses of UK Biobank have shown evidence of effects of BMI on social and socioeconomic outcomes in both Mendelian randomization and non-genetic within-sibling analyses (38). Here, we build on these previous analyses by including a broader set of social and socioeconomic outcomes, conducting additional sensitivity and secondary analyses, and facilitating comparisons across a range of health conditions and risk factors.

Higher genetic propensities towards asthma and eczema were estimated to reduce household income (mean difference = −£13,519, 95% CI: −£18,794 to −£8,243 for asthma, and mean difference = −£46,987, 95% CI: −£71,048 to −£22,925 for eczema). However, it is possible these estimates are susceptible to bias from pleiotropy, given the extreme size of the effects. Asthma and eczema share many genetic loci, along with inflammatory bowel disease and other autoimmune conditions (39). Therefore, the Mendelian randomization results for eczema and asthma may reflect an underlying genetic predisposition toward autoimmune condition susceptibility, rather than asthma or eczema specifically. This would not be detectable with Mendelian randomization sensitivity analyses if all SNPs included in the PRS were affecting autoimmune susceptibility rather than the conditions themselves (directional unbalanced pleiotropy). Additionally, the PRS for smoking initiation may capture impulsivity and risk taking as well as a propensity to smoke.

For some health conditions (asthma, breast cancer, eczema, migraine), we saw little evidence for observational (multivariable adjusted) associations with both socioeconomic and social outcomes, despite prior evidence often showing strong associations. For example, breast cancer has been associated with lower income (40), but there was no observational association between breast cancer and household income in UK Biobank. This could result from selection bias in UK Biobank (41), with participants potentially liable to have less severe/advanced forms of the condition or quicker recovery than all breast cancer patients across a population, and also to have greater financial support and better employment conditions than the general population. The effects of health conditions may also diminish over time; there is some evidence that the negative effect on income amongst breast cancer survivors reduces over time (40). It is therefore possible our study does not have the correct time frame to capture the effects of each health condition, or that well-functioning insurance markets and pension provision could mitigate socioeconomic effects of health conditions, at least within this generally affluent UK population (42). Additionally, if a participant developed any health condition after baseline, we would only know if the participant had a hospital episode which mentioned the condition.

There was evidence that depression was detrimental to multiple social outcomes, including reduced happiness, reported satisfaction rates, and increased loneliness. Given these are common features of depression this result was expected and gives us confidence the PRS for depression was suitably predictive of depression.

### Strengths and Limitations

The main strengths of this analysis are that Mendelian randomization analyses are generally less affected by confounding and reverse causation than multivariable adjusted (observational) analyses (43), and that UK Biobank is a very large sample with sufficient data to enable us to examine multiple health exposures and multiple socioeconomic and social outcomes. For some associations, there were marked differences between the Mendelian randomization and multivariable adjusted association estimates, which could result from reverse causation or confounding in the multivariable association adjusted estimates. For example, coronary heart disease was associated with a decreased chance of obtaining a university degree (APC = −8.1%, 95% CI: −9.0% to −7.1%) in the multivariable adjusted analysis, which is implausible given coronary heart disease usually occurs later in life than attending university, and this association was not seen in the Mendelian randomization analysis. Additionally, the SNPs contributing to the PRS were drawn from GWAS that excluded UK Biobank to avoid biases caused by sample overlap (36) and all reached genome-wide significance. Finally, the results from the main and split-sample analyses were largely consistent across exposures and outcomes, reducing the possibility of bias from differences in SNP effects between the GWAS and UK Biobank populations.

However, Mendelian randomization rests on assumptions that cannot be proven to be true (43). Assessing pleiotropy was difficult or impossible for many exposures, due to the low number of SNPs and wide CIs, but there was evidence for heterogeneity between SNPs for some associations (e.g. for income), and directional pleiotropy from Egger regression for a limited number of associations (e.g. for BMI on obtaining a university degree). As the outcomes were social and socioeconomic, not biological, the exclusion restriction assumption would be strong for any genetic variant (i.e. that the genetic variant affects the outcome only through the exposure). For example, we cannot assume that a SNP associated with income affects any health condition or risk factor solely through income. We therefore did not perform bi-directional Mendelian randomization (44), and so cannot rule out reverse causation for any analysis.

The PRS represent lifetime exposure to or risk for the health condition or risk factor, and interventions to reduce the exposure or risk of the exposure at different time points in a person’s life may have different effects; effects at specific points in life cannot be explored with the methodology used in this paper. As we used linear prediction models for all analyses, some effect estimates may also be impossibly large (i.e. over 100%), which could occur when precision is very low, though this was rare. Although Mendelian randomization is generally less affected by confounding and reverse causality than multivariable regression analyses, an important potential source of bias in these analyses is family-level effects. Recent evidence suggests that assortative mating and dynastic effects can lead to bias in Mendelian randomization effect estimates (45), with estimates of the effect of BMI on educational attainment being consistent with the null in within-family Mendelian randomization models using data from UK Biobank and the Norwegian HUNT study. In our previous analysis of UK Biobank (38), within-family Mendelian randomization models in UK Biobank alone were too imprecise to draw conclusions about whether the estimated effects of BMI on social and socioeconomic outcomes are robust to potential confounding by family-level factors. Since BMI is the exposure for which we have greatest statistical power (due to the strength of the genetic instrumental variable), we have not repeated the within-family analyses for our other exposures as power will be extremely limited. However, as more datasets are available that include genetic information for multiple family members, examination of whether these effects can be detected with a within-family Mendelian randomization design will be a high priority.

UK Biobank, while large, is not representative of the UK population as participants tend to be wealthier and healthier compared to the country as a whole, which may impart bias to our analyses (46). It is likely this biased some estimates towards the null, as wealthier and healthier people may be more resistant to any detrimental effects of health conditions and risk factors. Additionally, there is evidence of a geographic structure in the UK Biobank genotype data that cannot be accounted for using adjustment for principal components, which may also have biased our analyses (47). Some outcomes were dichotomised, which may have reduced our ability to detect associations (e.g. satisfaction with health).

For health conditions, the uncertainty around the Mendelian randomization effect estimates was large. As many health conditions had small associations with outcomes on multivariable adjusted analyses, this often meant the Mendelian randomization estimates were larger than the observed estimates or had a different sign, but this can be explained by the imprecision in the Mendelian randomization estimates. The uncertainty is due in part to the relatively poor ability of the PRS to predict some health conditions. There were minimal differences in prevalence between UK Biobank and the UK for most health conditions studied (apart from migraine and depression, which were less and more prevalent in UK Biobank respectively), but it is possible the health conditions were milder or better-managed in UK Biobank participants compared with the population as a whole (48). Therefore, null results should be interpreted as a lack of evidence for a causal effect, not evidence of a lack of a causal effect.

## 5. Conclusion

The results of this study imply that higher BMI, smoking and alcohol consumption are likely detrimental to socioeconomic outcomes. While the prevalence of smoking is decreasing in the UK (49), the average BMI has risen and is continuing to rise worldwide (50). Reducing average BMI levels, and further reducing smoking and alcohol intake, in addition to health benefits, may also improve socioeconomic outcomes for individuals and populations.

There was little evidence of causal effects of health conditions on socioeconomic outcomes, which may reflect true absence of causal effects or bias due to the characteristics of UK Biobank participants, or the low precision of our estimates for health condition effects.

## Data Availability

The empirical dataset will be archived with UK Biobank and made available to individuals who obtain the necessary permissions from the study’s data access committees. The code used to clean and analyse the data will be available here upon publication: https://github.com/sean-harrison-bristol/Effects-of-Health-Conditions-and-Risk-Factors-on-Socioeconomic-Outcomes

## Acknowledgements

LDH is funded by a Career Development Award from the UK Medical Research Council (MR/M020894/1). This work is part of a project entitled ‘social and economic consequences of health: causal inference methods and longitudinal, intergenerational data’, which is part of the Health Foundation’s Efficiency Research Programme. MG, SVK and DC work for the Health Foundation project entitled ‘Causal effects of alcohol and mental health problems on employment outcomes: Harnessing UK Biobank and linked administrative data’. The Health Foundation is an independent charity committed to bringing about better health and health care for people in the UK. The Medical Research Council (MRC) and the University of Bristol support the MRC Integrative Epidemiology Unit [MC_UU_12013/1, MC_UU_12013/9, MC_UU_00011/1]. The Economics and Social Research Council (ESRC) support NMD via a Future Research Leaders grant [ES/N000757/1]. PD acknowledges support from a MRC Skills Development Fellowship (MR/P014259/1). SVK acknowledges funding from a NHS Research Scotland Senior Clinical Fellowship (SCAF/15/02). The MRC/CSO Social & Public Health Sciences Unit, University of Glasgow is supported by the Medical Research Council (MC_UU_12017/13 & MC_UU_12017/15) and the Scottish Government Chief Scientist Office (SPHSU13 & SPHSU15). HEJ acknowledges support from an MRC Career Development Award in Biostatistics (MR/M014533/1). No funding body has influenced data collection, analysis or its interpretation. This publication is the work of the authors, who serve as the guarantors for the contents of this paper.

## Author Contributions

LDH, ARD, NMD, MD, HEJ and FR obtained funding for this study. SH cleaned and analysed the data and wrote the first draft. All authors contributed to study design, interpreted the results and revised the manuscript.

## Transparency Statement

Transparency statement: The lead author (the manuscript’s guarantor) affirms that this manuscript is an honest, accurate, and transparent account of the study being reported; that no important aspects of the study have been omitted; and that any discrepancies from the study as planned (and, if relevant, registered) have been explained.

---

**Supplementary Figure 1.**
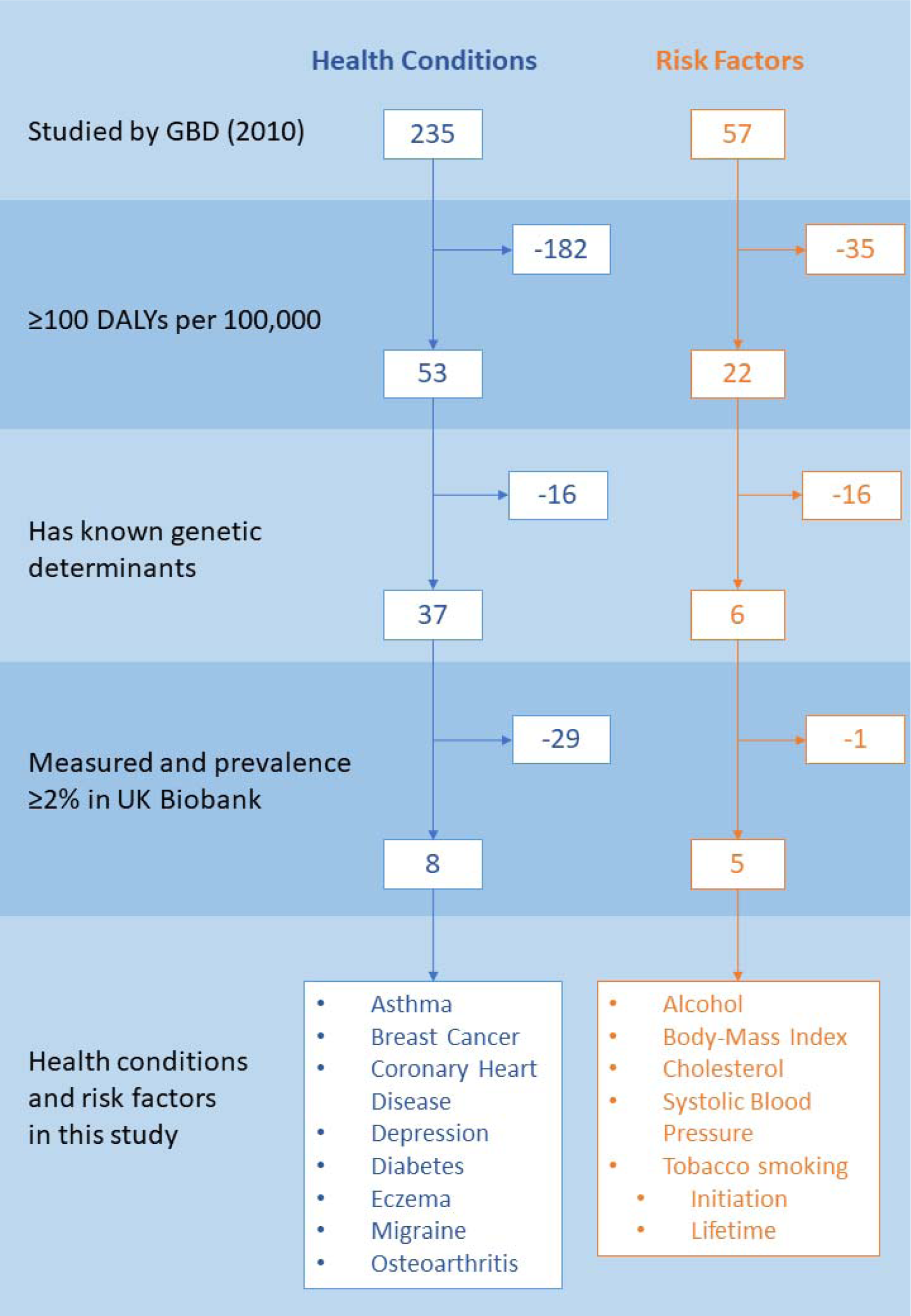
Flowchart of health conditions and risk factors

## References

1. Garland A, Jeon SH, Stepner M, Rotermann M, Fransoo R, Wunsch H, et al. Effects of cardiovascular and cerebrovascular health events on work and earnings: A population-based retrospective cohort study. Ann Intern Med. 2019;191(1):E3–10.

2. Hamood R, Hamood H, Merhasin I, Keinan-Boker L. Work Transitions in Breast Cancer Survivors and Effects on Quality of Life. Journal of Occupational Rehabilitation. 2018;1–14.

3. Wright C, Kipping R, Hickman M, Campbell R, Heron J. Effect of multiple risk behaviours in adolescence on educational attainment at age 16 years: A UK birth cohort study. BMJ Open. 2018;8(7).

4. Howe LD, Kanayalal R, Beaumont R, Davies AR, Frayling TM, Harrison S, et al. Effects of body mass index on relationship status, social contact, and socioeconomic position: Mendelian Randomization study in UK Biobank. bioRxiv [Internet]. 2019 Jan 1;524488. Available from: http://biorxiv.org/content/early/2019/01/18/524488.abstract

5. Garland A. Labor Market Outcomes: Expanding the List of Patient-centered Outcomes in Critical Care. Am J Respir Crit Care Med. 2017;196(8):946–7.

6. Johnson P, Stoye G, Sturrock D. Chief Medical Officer annual report 2018: better health within reach [Internet]. 2018. Available from: https://www.ifs.org.uk/publications/13786

7. de Leeuw E. Engagement of Sectors Other than Health in Integrated Health Governance, Policy, and Action. Annu Rev Public Health. 2017;38(1):329–49.

8. Wells M, Williams B, Firnigl D, Lang H, Coyle J, Kroll T, et al. Supporting “work-related goals” rather than “return to work” after cancer? A systematic review and meta-synthesis of 25 qualitative studies. Vol. 22, Psycho-Oncology. 2013. p. 1208–19.

9. Lawlor DA, Harbord RM, Sterne JAC, Timpson N, Smith GD. Mendelian randomization: Using genes as instruments for making causal inferences in epidemiology. Stat Med. 2008;27(8):1133–63.

10. Davey Smith G, Hemani G. Mendelian randomization: genetic anchors for causal inference in epidemiological studies. Hum Mol Genet [Internet]. 2014;23(R1):R89–98. Available from: http://www.ncbi.nlm.nih.gov/pubmed/25064373%5Cnhttp://www.pubmedcentral.nih.gov/articlerender.fcgi?artid=PMC4170722

11. Smith GD, Lawlor DA, Harbord R, Timpson N, Day I, Ebrahim S. Clustered environments and randomized genes: a fundamental distinction between conventional and genetic epidemiology. PLoS Med. 2007;4(12):1985–92.

12. Allen NE, Sudlow C, Peakman T, Collins R. UK biobank data: Come and get it. Science Translational Medicine. 2014;6(224).

13. Collins R. What makes UK Biobank special? Vol. 379, The Lancet. 2012. p. 1173–4.

14. Bycroft C, Freeman C, Petkova D, Band G, Elliott LT, Sharp K, et al. The UK Biobank resource with deep phenotyping and genomic data. Nature. 2018;562(7726):203–9.

15. Sudlow C, Gallacher J, Allen N, Beral V, Burton P, Danesh J, et al. UK Biobank: An Open Access Resource for Identifying the Causes of a Wide Range of Complex Diseases of Middle and Old Age. PLoS Med. 2015;12(3).

16. Murray CJL, Richards M a, Newton JN, Fenton K a, Anderson HR, Atkinson C, et al. UK health performance⍰: fi ndings of the Global Burden of Disease Study 2010. Lancet [Internet]. 2013;381(13):997–1020. Available from: http://dx.doi.org/10.1016/S0140-6736(13)60355-4%5Cnwhttp://www.ncbi.nlm.nih.gov/pubmed/23668584

17. Tyrrell J, Mulugeta A, Wood AR, Zhou A, Beaumont RN, Tuke MA, et al. Using genetics to understand the causal influence of higher BMI on depression. Int J Epidemiol [Internet]. 2018; Available from: https://academic.oup.com/ije/advance-article/doi/10.1093/ije/dyy223/5155677

18. Wootton RE, Richmond RC, Stuijfzand BG, Lawn RB, Sallis HM, Taylor GMJ, et al. Causal effects of lifetime smoking on risk for depression and schizophrenia: Evidence from a Mendelian randomisation study. bioRxiv. 2018;1–27.

19. Price AL, N.j. patterson, R.m. plenge, M.e. weinblatt, N.a. shadick. Principal components analysis corrects for stratification in genome-wide association studies. Nat Genet [Internet]. 2006;38(8):904–9. Available from: http://www.ncbi.nlm.nih.gov/pubmed/16862161

20. Gage SH, Munafò MR, Davey Smith G. Causal Inference in Developmental Origins of Health and Disease (DOHaD) Research. Annu Rev Psychol. 2016;67(1):567–85.

21. Craig P, Katikireddi SV, Leyland A, Popham F. Natural Experiments: An Overview of Methods, Approaches, and Contributions to Public Health Intervention Research. Annu Rev Public Health. 2017;38(1):39–56.

22. Tyrrell J, Jones SE, Beaumont R, Astley CM, Lovell R, Yaghootkar H, et al. Height, body mass index, and socioeconomic status: Mendelian randomisation study in UK Biobank. BMJ. 2016;352.

23. Kleibergen F, Paap R. Generalized reduced rank tests using the singular value decomposition. J Econom. 2006;133(1):97–126.

24. Harbord RM, Didelez V, Palmer TM, Meng S, Sterne JAC, Sheehan NA. Severity of bias of a simple estimator of the causal odds ratio in Mendelian randomization studies. Stat Med. 2013;32(7):1246–58.

25. Clarke PS, Windmeijer F. Instrumental variable estimators for binary outcomes. Vol. 107, Journal of the American Statistical Association. 2012. p. 1638–52.

26. Clarke PS, Windmeijer F. Identification of causal effects on binary outcomes using structural mean models. Biostatistics. 2010;11(4):756–70.

27. Burgess S, Labrecque JA. Mendelian randomization with a binary exposure variable: interpretation and presentation of causal estimates. Eur J Epidemiol. 2018;33(10):947–52.

28. Sterne JAC, Smith GD, Cox DR. Sifting the evidence—what’s wrong with significance tests? BMJ. 2001;322(7280):226.

29. Hayashi F. Econometrics. Princeton University Press. 2000. 233–234 p.

30. Haycock PC, Burgess S, Wade KH, Bowden J, Relton C, Smith GD. Best (but oft-forgotten) practices: The design, analysis, and interpretation of Mendelian randomization studies. Vol. 103, American Journal of Clinical Nutrition. 2016. p. 965–78.

31. Burgess S, Scott RA, Timpson NJ, Smith GD, Thompson SG. Using published data in Mendelian randomization: A blueprint for efficient identification of causal risk factors. Eur J Epidemiol. 2015;30(7):543–52.

32. Pierce BL, Burgess S. Efficient design for mendelian randomization studies: Subsample and 2-sample instrumental variable estimators. Am J Epidemiol. 2013;178(7):1177–84.

33. Greco M F Del, Minelli C, Sheehan NA, Thompson JR. Detecting pleiotropy in Mendelian randomisation studies with summary data and a continuous outcome. Stat Med. 2015;34(21):2926–40.

34. Hemani G, Bowden J, Davey Smith G. Evaluating the potential role of pleiotropy in Mendelian randomization studies. Hum Mol Genet. 2018;27(R2):R195–208.

35. Elsworth B, Mitchell R, Raistrick C, Paternoster L, Hemani G, Gaunt T. MRC IEU UK Biobank GWAS pipeline version 2. 2019.

36. Burgess S, Davies NM, Thompson SG. Bias due to participant overlap in two-sample Mendelian randomization. Genet Epidemiol. 2016;40(7):597–608.

37. Thorgeirsson TE, Geller F, Sulem P, Rafnar T, Wiste A, Magnusson KP, et al. A variant associated with nicotine dependence, lung cancer and peripheral arterial disease. Nature. 2008;452(7187):638–42.

38. Howe LD, Kanayalal R, Beaumont RN, Davies AR, Frayling T, Harrison S, et al. Effects of body mass index on relationship status, social contact, and socioeconomic position: Mendelian Randomization study in UK Biobank. Int J Epidemiol.

39. Paternoster L, Standl M, Waage J, Baurecht H, Hotze M, Strachan DP, et al. Multi-ancestry genome-wide association study of 21,000 cases and 95,000 controls identifies new risk loci for atopic dermatitis. Nat Genet. 2015;47(12):1449–56.

40. Jensen LS, Overgaard C, Bøggild H, Garne JP, Lund T, Overvad K, et al. The long-term financial consequences of breast cancer: A Danish registry-based cohort study. BMC Public Health. 2017;17(1).

41. Munafò MR, Tilling K, Taylor AE, Evans DM, Smith GD. Collider scope: When selection bias can substantially influence observed associations. Int J Epidemiol. 2018;47(1):226–35.

42. Fry A, Littlejohns TJ, Sudlow C, Doherty N, Adamska L, Sprosen T, et al. Comparison of Sociodemographic and Health-Related Characteristics of UK Biobank Participants with Those of the General Population. Am J Epidemiol. 2017;186(9):1026–34.

43. Davies NM, Holmes M V., Davey Smith G. Reading Mendelian randomisation studies: A guide, glossary, and checklist for clinicians. BMJ. 2018;362.

44. Zheng J, Baird D, Borges M-C, Bowden J, Hemani G, Haycock P, et al. Recent Developments in Mendelian Randomization Studies. Curr Epidemiol Reports. 2017;4(4):330–45.

45. Brumpton B, Sanderson E, Hartwig FP, Harrison S, Vie GÅ, Cho Y, et al. Within-family studies for Mendelian randomization: avoiding dynastic, assortative mating, and population stratification biases. bioRxiv [Internet]. 2019;602516. Available from: https://www.biorxiv.org/content/10.1101/602516v1?rss=1&utm_source=dlvr.it&utm_medium=twitter

46. Hughes RA, Davies NM, Smith GD, Tilling K. Selection bias in instrumental variable analyses. bioRxiv [Internet]. 2017;192237. Available from: https://www.biorxiv.org/content/early/2017/09/22/192237

47. Haworth S, Mitchell R, Corbin L, Wade KH, Dudding T, Budu-Aggrey A, et al. Common genetic variants and health outcomes appear geographically structured in the UK Biobank sample: Old concerns returning and their implications. bioRxiv [Internet]. 2018;294876. Available from: https://www.biorxiv.org/content/early/2018/04/11/294876

48. David Batty G, Gale C, Kivimaki M, Dreary I, Bell S. Generalisability of Results from UK Biobank: Comparison With a Pooling of 18 Cohort Studies. medRxiv (preprint) [Internet]. 2019; Available from: https://www.medrxiv.org/content/10.1101/19004705v1

49. Simpson CR, Hippisley-Cox J, Sheikh A. Trends in the epidemiology of smoking recorded in UK general practice. Br J Gen Pract. 2010;60(572):187–92.

50. NCD Risk Factor Collaboration, Lewington S, Clarke R, Qizilbash N, Peto R, Collins R, et al. Worldwide trends in body-mass index, underweight, overweight, and obesity from 1975 to 2016: A pooled analysis of 2416 population-based measurement studies in 128.9 million children, adolescents, and adults. Lancet. 2017;(2627–2642).

